# A resource efficient, high-dose neurorehabilitation program for chronic stroke at home

**DOI:** 10.1101/2024.10.08.24313178

**Authors:** Spencer A. Arbuckle, Anna Sophie Knill, Gabriela Rozanski, Michelle Chan-Cortés, Anastasia Elena Ford, Louis T. Derungs, David Putrino, Jenna Tosto-Mancuso, Meret Branscheidt

## Abstract

Accumulating evidence and medical guidelines recommend high-dose neurorehabilitation for recovery after stroke. Unfortunately, most patients receive a fraction of this dose, with therapist availability and costs of delivery being major implementational barriers. To explore a potential solution, we conducted a retrospective analysis of a real-world enhanced clinical service that used gamified self-training technologies at home under remote therapist supervision.

Data from 17 patients who completed a 12-18 week full-body, high-dose neurorehabilitation program entirely at home were analyzed. Program delivery relied on patients independently training (asynchronously) with the MindMotion GO gamified-therapy solution. Accompanying telerehabilitation sessions with a therapist occurred weekly while therapists used a web application to monitor and manage the program remotely.

Patients maintained high training adherence throughout and reached an average total Active Training Time—a measure more closely reflecting delivered versus scheduled dose—of 39.7±21.4 hours, with the majority (82.2±10.8%) delivered asynchronously. Patients improved in both upper-limb (Fugl-Meyer, +6.4±5.1; *p<*0.01) and gait and balance measures (Functional Gait Assessment, +3.1±2.6; *p<*0.01; Berg Balance Scale, +6.1±4.4; *p<*0.01). Most experienced subjective improvements in physical abilities and overall satisfaction. Per-patient therapist costs approximated 338 USD, representing a resource-efficient alternative to delivering the same dose in-person (1903 USD).

This work demonstrates effective high-dose neurorehabilitation delivery via gamified therapy technologies at home and shows that training time can be successfully decoupled from therapist-presence without compromising adherence, outcomes, or patient satisfaction. Given growing concerns over therapist availability and increasing health care costs, this resource-efficient approach can help achieve medical guidelines and complement existing clinic-based approaches.

## Introduction

Stroke is a critical global health issue, leaving many individuals with enduring disability (1) and placing an immense economic burden on healthcare systems. Rehabilitation is pivotal in fostering recovery and restoring functional independence. However, accumulating evidence suggests that significantly improving clinical outcomes hinges on the provision of high-dose training. Studies indicate that providing at least 20 hours of training over 10 weeks on top of standard therapy are needed to make a clinically meaningful impact (2–6). Therefore, the importance and clinical benefits of high-dose training are emphasized in existing and emerging medical guidelines for stroke rehabilitation. In the US, existing American Heart/Stroke Association guidelines recommend aerobic training of 60-300 minutes (over multiple sessions and days) per week in addition to strength and stretching exercises (7), while in the UK, the recent NICE guidelines update recommends at least 3 hours of targeted rehabilitation on 5 days per week (8).

However, despite strong evidence supporting the benefits of high-dose rehabilitation, most stroke patients fail to receive the recommended amount of therapy dose (6, 9–12). During acute inpatient care, patients were found to be active only 13% of their time and only 5.2% in contact with a therapist (13). A followup review nearly a decade later suggested little had changed (14). Furthermore, following discharge, patients complete less than 20 minutes of upper extremity training during approximately 45 minute sessions (15, 16). This issue becomes particularly pronounced in the chronic stage, where observational studies in outpatient settings have shown that overall training doses remain low: averaging just one purposeful leg or arm movement per minute (17, 18).

Thus, there is a stark gap between the evidence supporting high-dose neurorehabilitation versus what is delivered in traditional standard-of-care. Key implementational barriers that contribute towards this gap are human resource constraints (e.g., therapist cost and availability) and poor patient compliance and engagement (2, 4, 19, 20). Therapist availability in particular is only expected to worsen with a recent national US survey indicating that 66.6% of therapists intend to leave the profession within the coming year (21).

To address these barriers, numerous studies have tried to dissociate therapist time and outpatient infrastructure from therapy itself, either by incorporating technologies—particularly robotics—into in-clinic training programs (22); utilizing home-based training with or without the aid of additional technologies (23); incorporating a training model with a single therapist used to deliver care to multiple patients simultaneously (24, 25); or a combination of approaches (26). However, none of these approaches have fully overcome the implementational barriers: they often fail to free up therapist time, are costly, and have low patient adherence and engagement. To further complicate the matter, studies generally fail to demonstrate that scheduled training time equates to actual delivery of high-dose training—the vast majority do not report on actual therapy time delivered (27).

Thus, in this real-world enhanced clinical service and retrospective analysis, we investigated the extent to which implementational barriers related to delivering high-dose neurorehabilitation can be addressed with a technological approach. Our primary objective was to determine whether gamified training and monitoring technologies can be used to successfully deliver high-dose neurorehabilitation for chronic stroke survivors at-home without compromising either therapy adherence or clinical outcomes. By setting a soft target that 75% of the training dose was to be delivered through patient self-training (asynchronous) with the remaining 25% relying on synchronous telerehabilitation with the therapist, we attempted to dissociate therapeutic dose delivery from therapist availability.

Training was delivered with the aid of a gamified training technology that enabled full-body training of the upper-limbs, hand, trunk, and lower-limbs. Monitoring technology was employed to track actual training times and to ensure that therapists were able to consistently oversee patient progress and uphold the quality and success of the training. We performed separate evaluations of patient adherence, clinical effectiveness of training, patient satisfaction, and overall resource and cost efficiency of the program.

## Methods

### Patients

Chronic stroke patients were identified from the Mount Sinai Abilities Research Center Clinical Program registry. Patients were informed of the enhanced clinical service and offered the option to participate in high-dose neurorehabilitation (table 1). The program enrolled patients who were ≥22 years of age; had a first-time stroke ≥3 months prior; presented with either arm weakness and/or difficulty with balance; were able to follow multi-step commands; and showed willingness and ability to commit to the program length and the weekly training dose accompanied with virtual visits. Patients inappropriate for the program included those with plegia of the affected upper limb; severe pain; or severe cognitive or physical challenges which made it difficult for them to participate in neurorehabilitation.

**Table 1.** Baseline characteristics of analyzed patients.

|  |  |
| --- | --- |
| n | 17 |
| Age, y | $54.8 \pm 14.1$ |
| Time post stroke, y | $5.4 \pm 4.7$ |
| Sex |  |
| Men | 9 |
| Women | 8 |
| Stroke Type |  |
| Ischemic | 9 |
| Hemorrhagic | 8 |
| Affected Hemisphere |  |
| Left | 6 |
| Right | 11 |

Twenty-six patients were enrolled into the program: 23 patients started training (supp. fig. S1) of which 6 either dropped out or were lost to follow-up. Dropout reasons included changes in medical status and refusal to partake in in-person discharge assessments. 17 patients completed the training program (8 women; mean age = 54.8 ±14.1 years; time post-stroke 5.4 ±4.7 years), and their data was analyzed retrospectively. There were no significant differences regarding chronicity, age, or impairment (tested with the baseline Fugl-Meyer Upper Extremity and Berg Balance Scale measures) between patients who completed the training program and those who did not, and no adverse events were reported during the program.

All data was collected and stored in a data registry approved by the Mount Sinai Program for Protection of Human Subjects institutional review board (Icahn School of Medicine at Mount Sinai, New York, U.S.A.; STUDY-21-00345). All data analysis was conducted in adherence with the Declaration of Helsinki and the local legislation and institutional requirements.

### High-Dose Telerehabilitation Training Program

All patients were assigned to a single-arm treatment group involving a high-dose, home-based training program (fig. 1A-B). Overall, the program goal was to deliver 36 hours of training during the program. The target dose was consistent with a previous large telerehabilitation study that showed significant improvements in upper-limb impairment (42 hours; see (28)). In contrast, training in this program was not limited to one body area (29–31) and could be achieved with multiple effectors (i.e., upper-limb, hand, trunk, and lower-limb).

**Fig. 1.**
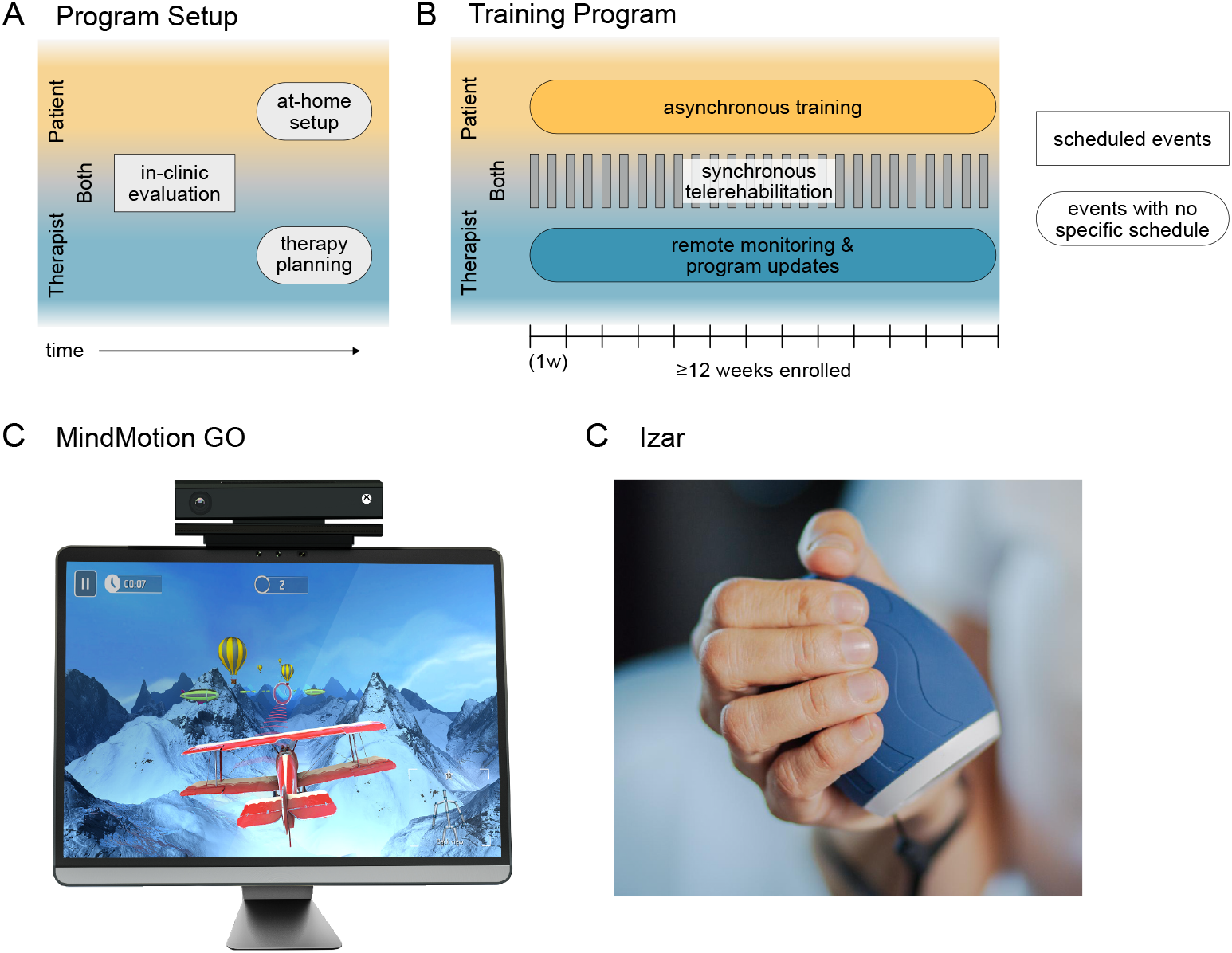
Design and technologies of the home-based training program. Patients were enrolled into a home-based training program that consisted of self-directed (asynchronous) training sessions performed by the patient each week complimented by multiple weekly (synchronous) sessions directly supervised by a physical therapist. (**A**) Prior to program start, patients were evaluated in-clinic by a physical therapist. The therapist would then create a training program for the evaluated patient. Patients set up the training system (MindMotion GO and Izar) at home. (**B**) During the training program, synchronous telerehabilitation sessions were scheduled as per therapist and patient availability, whereas asynchronous training (done by the patient) or remote monitoring (done by the therapist) could be completed at any time. Patients additionally underwent an in-clinic assessment battery prior to the program starting and upon program completion to evaluate clinical efficacy of the training program (not shown). (**C**) The MindMotion GO telerehabilitation device uses camera-based motion tracking to control gamified training activities displayed on a TV-screen. (**D**) The Izar, a therapeutic device to train hand function, being grasped in a hand.

Patients were given the freedom of achieving the 36 hours of training dose over a period between 12 and 18 weeks and were given a high-level goal to train for at least 120 minutes per week. Consistent with in person rehabilitation practice, patients could take short breaks away from the program (these breaks were incorporated in the analysis; see *Methods - Binning training data into weeks*). Patients were also free to continue additional regular rehabilitation services. Dose was delivered either via synchronous telerehabilitation or asynchronous training (table 2).

**Table 2.** Suggested weekly schedule.

|  | Planned session duration (min) | Planned sessions per week (#) | Expected training per session (min) | Expected training per week (min) |
| --- | --- | --- | --- | --- |
| Asynchronous training | at patient discretion | at patient discretion | at patient discretion | $\geq 90$ |
| Synchronous training | 30 | 1-3 | 15 | 30-45 |
| Total | | | | $\geq 120$ |

### Synchronous sessions

These telerehabilitation sessions were guided directly by a physical therapist who was present virtually. The therapist was free to schedule 30-minute synchronous sessions with a patient at a frequency determined by their clinical judgment. The content of the sessions included gamified training using the MindMotion GO system (see *Methods - Full-body gamified system for asynchronous training*) under visual observation of the therapist and check-in conversations between the patient and the therapist to discuss training goals, training adjustments, feedback, etc.

#### Asynchronous sessions

These were self-directed training sessions where patients performed gamified training in a remote environment (e.g., at home). In contrast to synchronous training, asynchronous sessions did not include a therapist present. Patients had the ability to independently choose when and for how long to train asynchronously, and the ability to skip or repeat prescribed activities. At least 75% of a patient’s total dose was expected to be delivered asynchronously.

Asynchronous training content for each patient was determined by their supervising therapist, who tailored their training program by selecting gamified activities on the MindMotion GO based on the patient’s disease severity, therapeutic goals, preferences, and their physical capacity, with the range of motion calibrations defining the playable range of each activity. This calibration ensured that the gameplay was within the patient’s safety envelope (to minimize pain and adverse events such as risk of falls), and was updated as needed. The gamified activities targeted one or more areas of the following: hand, upper limb(s), trunk, and/or lower limb(s).

#### Full-body gamified system for asynchronous training

A commercially available technology (MindMotion GO; FDA Class-II) was used to deliver full-body gamified training. The MindMotion GO is a real-time, motion capture-based neurorehabilitation therapy technology (fig. 1C) that *i)* enabled therapists to set up and track training, *ii)* enabled patients to perform asynchronous training sessions, *iii)* provided teleconsultation capabilities for synchronous training and check-ins, and *iv)* captured performance measures during gameplay that allowed for the quantification of dose and nature of training delivered to each patient. At the time of this enhanced clinical service, the MindMotion GO catalog included 36 gamified activities designed to train specific movements relevant to neurorehabilitative therapy (see supp. table S1).

Patients’ body and limb kinematics were captured by accompanying optical hardware sensors (Kinect, LeapMotion), which were used to drive mechanics of the gamified activities. An additional peripheral device (Izar; fig. 1D) was used to capture fine-grained forces generated during hand and wrist control and use them for gameplay.

A range of metrics related to a patients’ training performance was captured in real-time and uploaded to a web platform (Companion). Therapists used this web platform to remotely set up or adjust therapy plans and to track training progress.

Equipment was shipped to a patient’s home, where it was connected to a TV. All but one patient successfully installed the system without requiring in-person assistance from the therapy team. Customer and technical support (via telephone or video-call) was provided for the duration of the program.

#### Quantification of dose and nature of training

We analyzed training data from the MindMotion GO device to assess program feasibility and patient behavior, including training time, duration, and movements trained.

#### Quantifying dose delivered

*Active Training Time* (ATT), defined as the time spent engaging in gamified activities (excluding setup, calibration, or pauses), was used as a measure of training dose delivered.

#### Binning training data into weeks

Our primary focus was on training program dosage (i.e., hours of training) rather than program duration (i.e., weeks in the program). However, to investigate training behavior over time, training data were organized into *training weeks*, defined as consecutive 7-day periods starting on the patient’s program start date and ending at their exit assessment. Training weeks could vary across patients and if a patient missed training in a week (i.e., zero ATT), it was excluded (5.8% of all total 346 enrolled weeks). For clarity, we use *trained weeks* in the figures (fig. 2A & C), acknowledging that these may not reflect consecutive weeks.

**Fig. 2.**
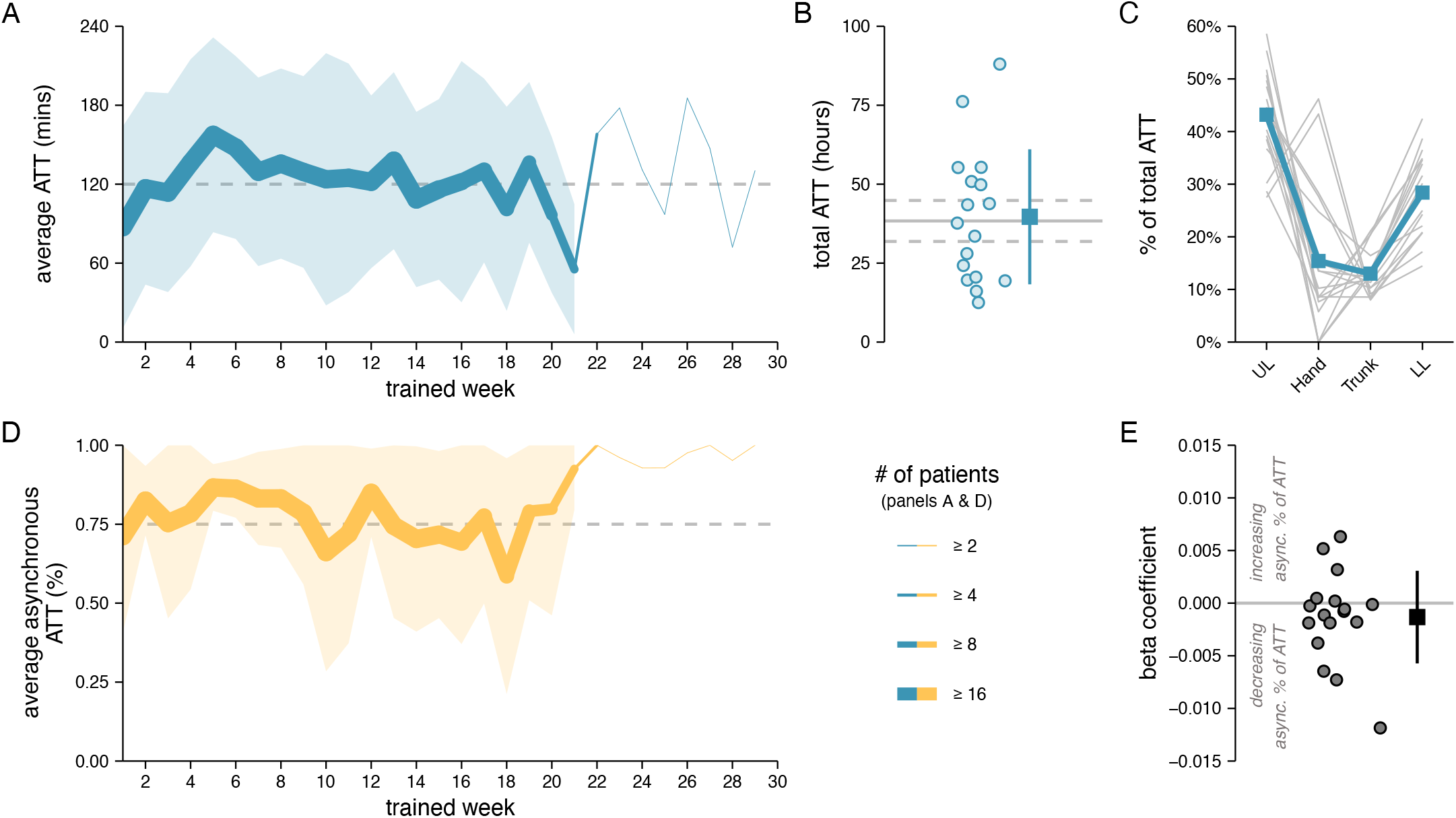
Training dose. (**A**) Active training time (ATT) per week averaged across patients. The thickness of the line represents the number of patients who remained in the program by that training week. The shaded area indicates the standard deviation above and below the average (for weeks with n*>*3), and the dashed line indicates the target weekly dosage of 120 minutes. (**B**) The total ATT dosage achieved at program end. Each dot corresponds to one patient. The blue box indicates the average and the vertical line indicates the standard deviation above and below the average. The solid gray line indicates the total expected dosage if each patient achieved 120 minutes ATT per week they were training in the program, with the dashed lines reflecting the standard deviation around the average expected dose (patients completed a varying number of training weeks in the program). (**C**) Proportion of total ATT that patients spent training their upper limbs (UL), hands, trunk, or lower limbs (LL). Each patient’s data is plotted as a gray line with the average in blue. (**D**) The proportion of weekly ATT that was achieved asynchronously, averaged across patients. Similar formatting as in **A**. (**E**) Quantifying patients’ tendency to increase, decrease, or maintain their ratio of asynchronous-to-synchronous ATT across training weeks (see Methods). On average, patients maintained consistency across weeks in the proportion of ATT achieved asynchronously.

#### Assessing training consistency

We examined whether patients’ asynchronous training behavior changed over time using ordinary least squares regression. A positive or negative beta coefficient indicated whether the proportion of ATT trained asynchronously tended to increase or decrease, respectively, across weeks. The magnitude of a coefficient represents the weekly change rate (e.g., a coefficient of 0.001 is +1% increase per week). The beta coefficient for each patient is plotted in Figure 2E.

#### Analyzing training schedules

We tested if patients had idiosyncratic but nevertheless reliable training schedules. To assess patients’ training schedules, we calculated the distribution of their total asynchronous ATT across each hour of each day of the calendar week. For each patient, data from even- and odd-numbered training weeks were binned separately. We then computed Pearson’s correlation between training schedules for even- and odd-numbered weeks within patients to measure schedule reliability. To quantify the uniqueness of each patient’s schedule across the cohort, we computed the average Pearson’s correlations between their even- and odd-week schedules and those of all other patients’ schedules. The within-patient and across-patient correlations are plotted in Figure 3E.

**Fig. 3.**
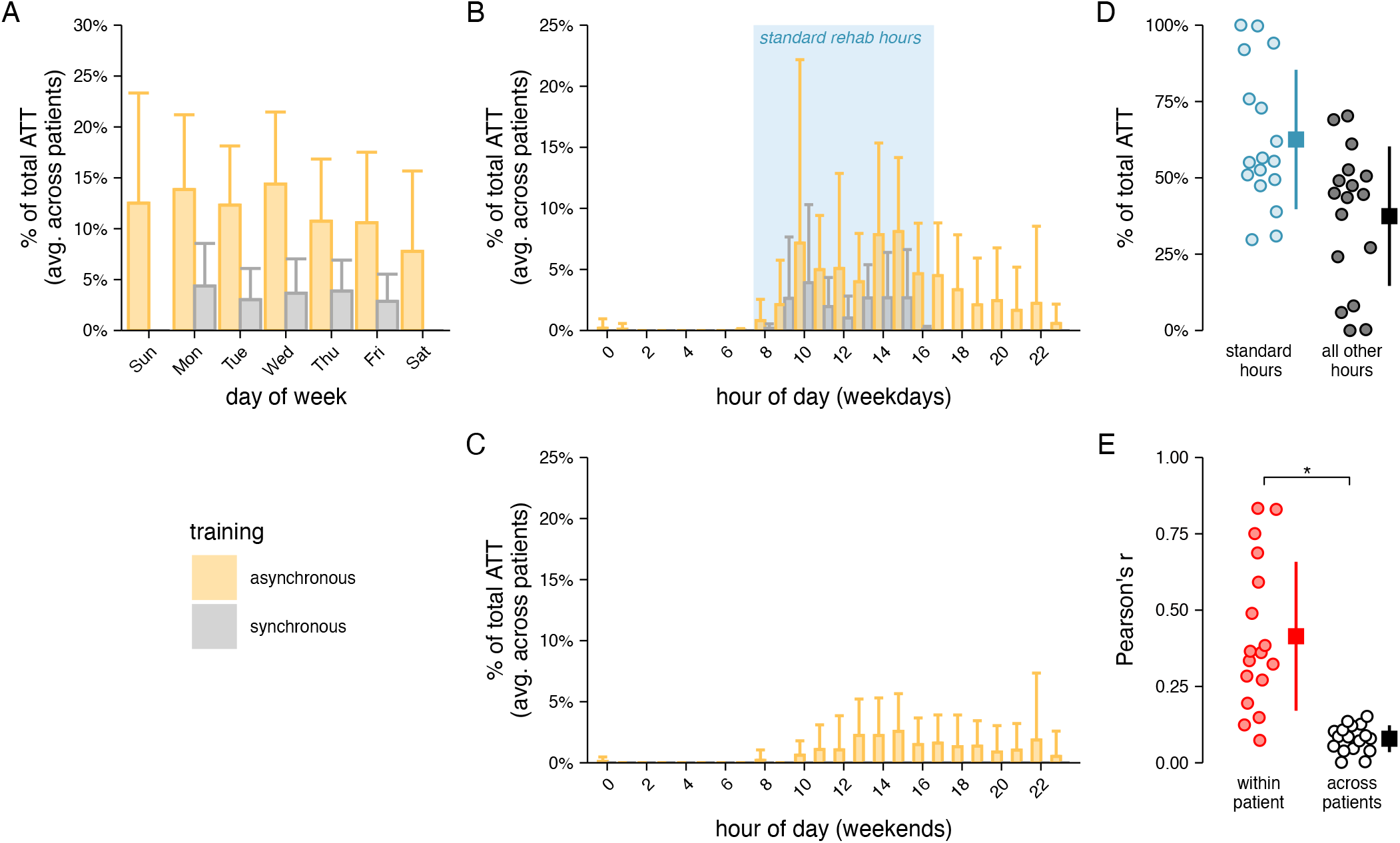
Training distribution across days of the week and times of the day. (**A**) Bar chart depicting the proportion of total ATT (averaged across patients) binned according to the day of the week. Synchronous training times were only logged during the weekdays. Error bars reflect the standard deviation across patients. (**B-C**) Bar charts depicting the proportion of total ATT (averaged across patients) binned according to the starting hour of the training session for training done on weekdays (**B**) and weekends (**C**). Similar formatting as in **A**. (**D**) Percent of total ATT that occurred (blue) / did not occur (gray) during standard working hours, when the therapist was available for synchronous training sessions (blue box in **B**). Each dot corresponds to one patient. The boxes indicate the averages and the vertical lines indicate the standard deviations above and below the averages. (**E**) Quantifying uniqueness of patients’ average weekly asynchronous training profiles (see *Methods*). Each dot in the within-patient grouping (red) reflects a patient’s consistency of their weekly training profile, and each dot in the across patients grouping (white) reflects the average correlation between that patient’s training profile and the training profiles of all other patients. The boxes and vertical lines are formatted the same as in D. *significant difference between within patient and across patient correlations (two-sided paired *t* test, *p<*0.05).

### Capturing clinical outcomes and patient subjective experiences

We quantified the effect of training on patients’ impairment and function. As training was delivered full-body, we captured a wide range of functional outcome measures and patient reported outcome measures (PROMs) to quantify training-related changes across multiple effectors. A comprehensive list of all outcome measures is available in the supplementary material (supp. tables S2 and S3).

Clinical outcome measures and PROMs were captured at the beginning and the end of the program. The clinical outcome measures were assessed in person at the Abilities Research Center by a physical therapist who was not their supervising therapist to retain blindness. PROMs were completed by patients through online surveys captured via REDCap electronic data capture tool (32, 33). A custom survey was administered to all enrolled patients at the end of the program, regardless of training completion, to gather information on their experiences, satisfaction, motivation, and perceived improvements.

### Cost analysis

We conducted a simplified cost analysis, comparing physical therapist resourcing costs for two scenarios: *i)* this program, with training delivered via a combination of asynchronous and synchronous sessions, and *ii)* a scenario where the entire dose is delivered through direct supervision. The median hourly salary for a physical therapist was sourced from the US Bureau of Labor Statistics 2023(34), which excludes fringe benefits (e.g., medical insurance). Therapist costs for both scenarios were calculated by multiplying the total training hours by the median hourly salary (in USD), adjusted for the percentage of training delivered under supervision.

### Statistical analysis

All analyses and statistical tests were performed using R (version 4.3.1; 35–38). Values presented in the results are the average across patients (plus or minus the standard deviation). For Pearson’s correlation values, we first Fisher z-transformed the correlations and calculated the mean and standard deviation. The mean and standard deviation were then transformed back to correlations and reported according to convention. To compare if the proportion of asynchronous ATT systematically changed for all patients across the program duration, we used a two-sided *t* test to compare the beta coefficients to zero (see *Methods - Assessing training consistency*). To compare if patients had idiosyncratic asynchronous training schedules, we used a two-sided paired *t* test to compare the Fisher z-transformed correlations from within and across-patient comparisons of their training schedules (see *Methods - Analyzing training schedules*). We set the significance level to *p* = 0.05 for these statistical tests. For the analysis of clinical improvements, changes from baseline to program end were tested using paired two-sided paired *t* tests. As these tests involved multiple comparisons without any specified *a-priori* hypotheses, we adjusted the significance level by dividing by the number of clinical outcomes examined which was 12 (i.e., Bonferroni correction). For clarity, we report uncorrected *p* values in the text. Note that not all 17 patients were evaluated on all assessments and therefore, the degrees of freedom varied across the clinical outcomes.

Results of the exit survey are reported as a proportion of total survey respondents, which differs from patients who completed the program. We chose to present the exit survey results for all respondents to ensure all views were included, irrespective of whether a patient officially completed the program. Nineteen patients completed the exit survey, of which 16 completed the program and 3 did not (2 dropped out, 1 was lost to follow up).

## Results

### Long-term, high-dose training can be achieved for chronic stroke patients in the home

A retrospective analysis was carried out on 17 chronic stroke patients who completed high-dose neurorehabilitation. Overall, the goal was for patients to receive a minimum 36 hours of Active Training Time (ATT; see *Methods*), delivered entirely at home and spread across all major motor functions (i.e., upper-limbs, hand, trunk, and lower-limbs). Training was delivered using a gamified therapy solution (MindMotion GO with Izar), through a combination of the patients following prescribed training plans on their own (self-directed asynchronous training) supplemented by weekly training sessions with a therapist being virtually present (therapist-directed synchronous telerehabilitation). Therapists were able to remotely define and alter training plans and maintain oversight of the program using a web-based application for patient monitoring (Companion).

Training duration and consistency was high. Patients trained an average of 120.51 ±52.47 minutes per week for 19.18 ±3.24 weeks (fig. 2A), with all patients exceeding the intended minimum duration of 12 weeks. Overall, patients achieved an average cumulative training dose of 39.67 ±21.37 hours, slightly surpassing the program goal of 2 hours of ATT per week (fig. 2B). This cumulative training dose was full body, with a slight tendency towards upper limb training (43.20 ±9.06% of total ATT) compared to the lower limb (28.40 ±8.27%), hand (15.40 ±14.09%) and trunk (13.01 ±4.25%; see fig. 2C). Altogether, these results demonstrate the feasibility of asynchronous/synchronous programs to deliver actual high-dose training distributed across the full-body.

It is important to note that the ATT dose measure used in this analysis is a closer representation of dose actually delivered in an intervention, as compared to the scheduled dose that is almost exclusively reported in other high-dose neurorehabilitation studies (5, 10, 28, 39, 40). Furthermore, this dose was delivered in addition to any other rehabilitation received by patients. Indeed, 11 out of the 17 patients reported also receiving care outside the program (e.g., physiotherapy, occupational therapy, or general exercise), highlighting the program’s complementarity to standard of care neurorehabilitation.

Despite the extended program duration of approximately 19 weeks, patients reported high levels of satisfaction. In the exit survey, 73.7% of respondents indicated they were either very satisfied or satisfied with the program (supp. fig. S2A). Additionally, patients expressed strong motivation to continue training beyond the program length, with 57.9% reporting they were very motivated or motivated to continue (supp. fig. S2B). This was particularly apparent in one patient who trained for 29 weeks, far surpassing the official program length.

### Majority of program dose was delivered without a therapist present

Therapists scheduled synchronous telerehabilitation sessions with patients as per their clinical discretion to ensure compliance to the overall program. However, the goal was for patients to achieve much of their training dose asynchronously. Of the cumulative dosage trained by patients, approximately 34.18 ±21.72 hours were delivered asynchronously. Per patient, this corresponded to 82.19 ±10.81% of their total ATT (fig. 2D), constituting the majority of their overall dose and higher than the 75% target.

Patients continued to complete most of their training asynchronously as the program progressed (fig. 2D), with no systematic change in the ratio between synchronous and asynchronous training dose from week-to-week (fig. 2E; average 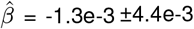, two-sided *t* test vs. 0: *t* 16 = -1.24, *p* = 0.23).

All synchronous sessions were delivered during regular 8am-5pm (Mon-Fri) working hours (fig. 3A-C). In contrast, asynchronous training offered patients the flexibility of training at any time, uncoupled by restrictions of therapist availability. Indeed, a substantial proportion of total ATT (37.45% ±22.84%, 15.70 ±12.29 hours) was logged outside of working hours and on weekends (fig. 3D).

Patients’ own schedules imposed restrictions on when they were able to train. Interestingly, while the majority of patients remained relatively consistent in their overall training schedules from week-to-week (within patient Pearson’s *r* = 0.46 ±0.34, red dots in fig. 3E), the training schedules were highly individual (across patients Pearson’s *r* = 0.08 ±0.04, white dots in fig. 3E; two-sided paired *t* test vs. within-patient schedule reliability: *t* 16 = -4.74, *p* = 2.22e-4).

Taken together, these results demonstrate that patients remained highly adherent to the home-based training program even if training was largely delivered asynchronously.

### Patients showed significant improvements in gait, balance, and upper-limb function

Next, we investigated the effect of the high-dose training on patients’ recovery and self-reported wellbeing. As the delivered training was full-body, we captured a wide range of standardized upper-limb, gait and balance, and physiological measures to quantify training related changes in impairment and function across multiple effectors (see *Methods*). All measures were obtained at the beginning and end of the training program.

Patients demonstrated positive improvements in the three core impairment and functional measures for multiple effectors (see table 3)namely the Fugl-Meyer Upper Extremity assessment for upper limb impairment (FM-UE; +6.41 ±5.09, *n* = 17, *p* = 8.8e-5), the Berg Balance Scale for generalized balance and transfer stability (BBS; +6.07 ±4.43, *n* = 15, *p* = 2.5e-2), and the Functional Gait assessment (FGA; +3.07 ±2.55, *n* = 15, *p* = 3.7e-4) for postural stability and walking. Amongst these, both the FM-UE and BBS were above the Minimal Detectable Change (MDC) for chronic stroke (MDC = 3.2 and 4.66 points respectively; see 41, 42), whereas the FGA was not (MDC = 4.2 points; see 43). Although there were positive improvements across most other captured measures, they did not cross thresholds for either statistical significance or minimum detectable change (see supp. table S2). These positive improvements in standard clinical measures were complemented by 63% of respondents self-reporting subjective improvements in their physical abilities and well-being (supp. fig. S2C).

**Table 3.**
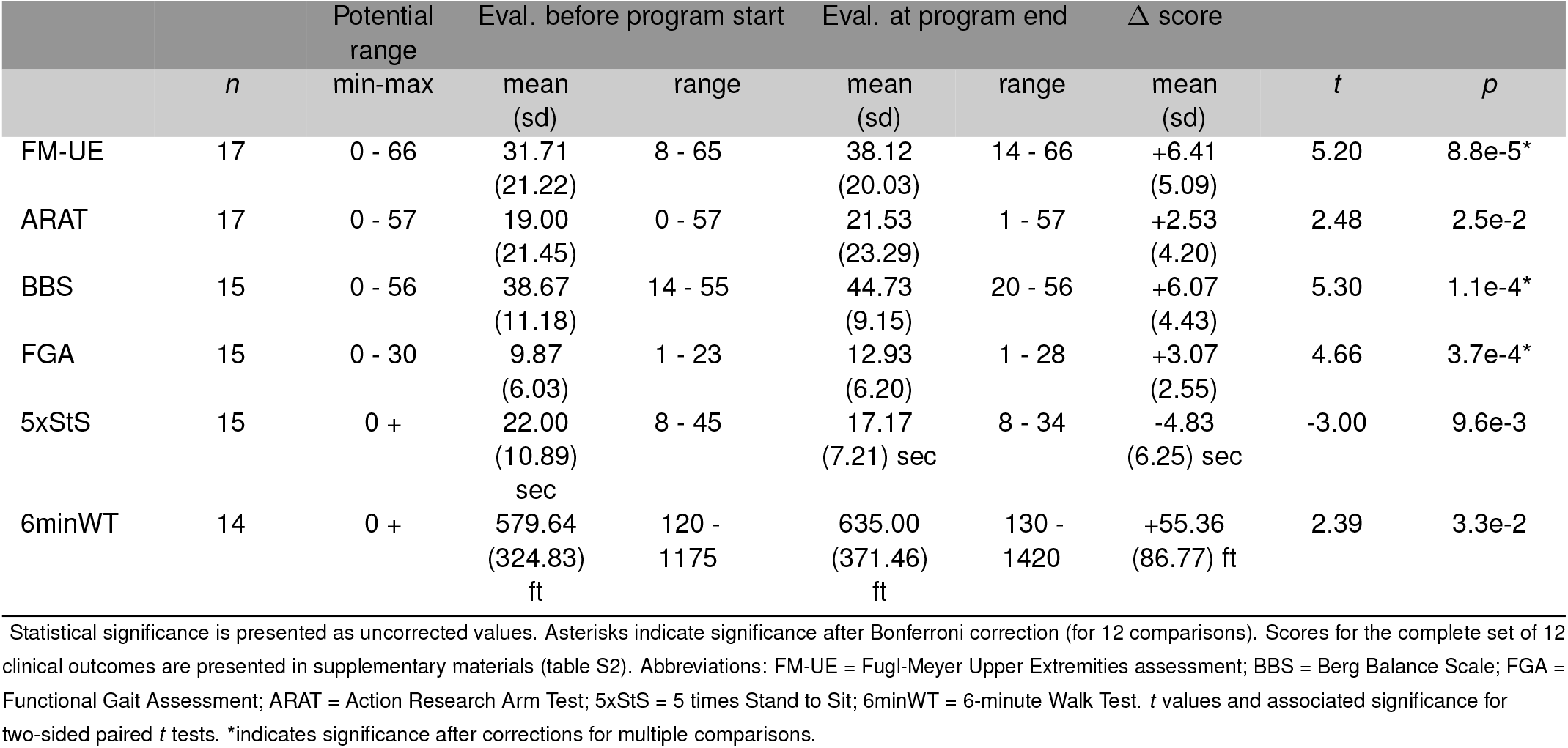
Core clinical outcomes.

Overall, these results demonstrate that patients receiving a high-dose training program at home showed significant improvements in outcome measures related to full-body impairment and function, with these improvements reinforced by patients’ self-reported measures.

### Asynchronous training significantly reduces costs associated with delivery of high-dose training

Finally, we computed the personnel costs associated with the delivery of this high-dose training. Based on the current analysis, 39.7 hours of cumulative training dose was delivered to each patient (on average), with the majority being delivered asynchronously (82.2%). Therefore, direct therapist presence was required to deliver only 17.8% of the training dose, and thus the total therapist costs per patient would

Potential Eval. before program start Eval. at program end Δ score amount to 338 USD (median hourly salary for physical therapist = 47.94 USD; see 34).

In contrast, the delivery of regular therapy hours relies on direct therapist presence either through in-clinic visits or using teleconferencing tools (44). Therefore, under a scenario where all training is delivered directly, the costs of delivering 39.7 hours would amount to 1903 USD—a substantial increase of 1565 USD when compared to the current hybrid asynchronous / synchronous program.

## Discussion

While the evidence for high-dose training in stroke rehabilitation is accumulating, the real challenge is in its implementation: how to deliver high-dose training effectively and in a resource-efficient way? Here we demonstrate that a gamified neurorehabilitation program can be used to successfully deliver high-dose full-body training to chronic stroke patients at home. Using a combination of therapist-directed telerehabilitation sessions (synchronous) and patients’ training on their own (asynchronous training), we were able to achieve resource-efficient dose delivery—the program required only a fraction of the therapist’s time compared to traditional one-to-one in-person neurorehabilitation sessions. To our knowledge, this work is the first to demonstrate a significant decoupling of active training dose delivery from therapist presence.

Critically, however, this decoupling was not at the expense of clinical outcomes. Unlike most previous approaches, patients showed positive improvements in all measures related to gait, balance, and upper-limb function, thus addressing that stroke typically results in multi-limb deficits. These improvements were noticeable by patients, with 63% reporting higher physical abilities and overall well-being.

### The role of gamified technology to deliver high-dose training at home

A critical issue in most neurorehabilitation studies is the lack of transparent reporting between scheduled therapy and what is truly delivered. A recent review identified the scale of the problem, with approximately only 13% of studies reporting on the actual training dose received (27). When regular therapy is quantified, the actual time on task is a fraction of that scheduled, and is significantly lower than training doses required for recovery (45).

Using technologies has two advantages here: *i)* namely as a means of sensitively and accurately quantifying the amount of training dose delivered, while *ii)* also providing a set of training activities that homogenizes the nature of training received by the patient. Such technologies therefore respond to a growing demand for better documentation and transparency of dose delivery in neurorehabilitation (46).

A key element of this program was the significant use of asynchronous training delivery to achieve higher dose. Our core mitigation strategy against reductions in patient engagement and adherence (47) was to ensure that the therapist was always in-the-loop, with the synchronous telerehabilitation sessions serving as a means to review and update plan of care, but also to ensure patient engagement. Additionally, gamified activities, a wide variety of exercises, and user-friendly interfaces were identified as important components (48) and were implemented in the technology used. Our results demonstrate the success of these mitigation strategies, with patients able to self-administer over 80% of the total training dose and maintain consistent engagement.

That patients remained engaged in the program suggests they could realize a principal advantage of training asynchronously: convenience. Patients were able to train anytime without being restricted by therapist availability. Indeed, here we found that when patients were given a high-level training goal, supplied with tailored training programs delivered through gamified technologies, and provided with therapist check-ins, they utilized both weekdays and weekends to train, which is not possible with conventional inperson neurorehabilitation. Furthermore, patients tended to train with highly idiosyncratic schedules that were most likely driven by unique work, life, and social commitments.

### Comparing approaches to deliver high-dose training

In previous studies, a variety of different approaches for delivering high-dose training have been investigated, but few are truly scalable in a resource-constrained healthcare landscape. High-dose approaches that require one-on-one in-clinic therapy sessions are prohibitively expensive, are often limited to urban facilities, and struggle to scale (i.e., treat more patients) without significant staffing (5). Offering one-to-many training can mitigate the staffing and cost concerns, but often impose restrictions on patient scheduling. Supplementing regular therapy with robotic systems imposes high equipment and maintenance costs (49), and the complexity of these devices constrains them to supervised use in-clinic (50).

In comparison, while home-based programs are an attractive alternative, they are not all equal. Programs that rely solely on passive exercise videos may be inexpensive, but they often struggle with poor therapy adherence rates and a limited ability to customize care plans. And while traditional telerehabilitation benefits from direct therapist and patient contact with respect to adherence, it is again dependent on therapist availability (47).

What remains attractive and potentially scalable, however, are home-based programs that use a hybrid combination of asynchronous and synchronous training. By using gamified and monitoring technologies to enable asynchronous training, these programs are able to overcome many of the known challenges for the approaches described above.

Recent studies have demonstrated the efficacy of providing high-dose training at home (28, 29), however the full value of these programs were not realized; there were significant requirements on therapist availability (50:50 synchronous/asynchronous split) and no accompanying cost analysis. The work here extends these results and demonstrates that technology-enabled asynchronous training allows for high training doses to be delivered with limited demands on therapist availability (82% was delivered asynchronously). Despite limited therapist presence in the program, patient engagement and satisfaction remained high. We also report costs associated with program delivery which represents a fraction of the resource costs associated with delivering equivalent high-dose training through scaling up one-to-one neurorehabilitation.

### Integrating asynchronous training into standard-of-care

For home-based neurorehabilitation programs to gain widespread adoption, it is critical that they integrate into and complement current neurorehabilitation workflows. Here we offer two pragmatic suggestions. For patients that routinely receive in-person outpatient rehabilitation, part of the in-person sessions can be used to review, discuss, and update the plan of care for remote asynchronous training. In contrast, for patients in more rural settings, in-person visits can be reduced in lieu of synchronous telerehabilitation training, again complemented by training asynchronously. Indeed, following the COVID pandemic, synchronous telerehabilitation in the US remains a billable service due to public health emergency extensions (51), while new Current Procedural Codes (CPT) billing codes have been established by the Center of Medicare & Medicaid Services to provide reimbursement pathways for technologies that enable asynchronous neurorehabilitation services (52, 53).

Despite the attractiveness of a high-dose neurorehabilitation at home, a few caveats should be considered. In such programs, there is a tangible concern that the importance of therapists are reduced. We unequivocally disagree with this notion. Therapists are and will remain essential for building collaborative relationships with patients that are critical for engagement and recovery (54).

To this end, it is essential that the technologies are designed with therapists’ needs in mind. Care should also be taken so that therapists do not suffer from fatigue and burnout due to inherent demands of teleconferencing calls (55).

Finally, for patients, although home-based asynchronous training offers significant benefits, it may not be suitable for everyone. Some patients have historically preferred in-person therapy (56, 57). Furthermore, challenges including limited physical space, privacy concerns, and the need for technical proficiency are major barriers to widespread adoption of training technology (56). With respect to this last item, the relatively young average age of our cohort might suggest higher technical literacy and fewer cognitive impairments. Deficits in vision, communication, cognition, or a heightened fall risk may also significantly impact the effectiveness of predominantly asynchronous therapy programs and raise safety concerns (58, 59). Overall, these points only serve to highlight the critical role of therapists in properly selecting and recommending home-based neurorehabilitation only to appropriate patients.

## Conclusion

This work introduces a scalable model for delivering high-dose training to stroke patients, effectively addressing therapist shortages with an asynchronous and gamified technology-driven approach. This work, to our knowledge, is the first demonstration of the full potential value of hybrid asynchronous and synchronous high-dose neurorehabilitation programs at home. The program described here is potentially an attractive option to bridge the gap between accumulating scientific evidence and medical guidelines around the need for high-dose training, and the reality of traditional neurorehabilitation today.

## Data Availability

Aggregated and de-identified data that support the findings of this study are available from the corresponding authors upon reasonable request.

## Supplementary material

Accompanying supplemental material is available on the medRxiv preprint server.

