## Supplemental material for "A resource efficient, high-dose neurorehabilitation program for chronic stroke at home"

Supplementary material

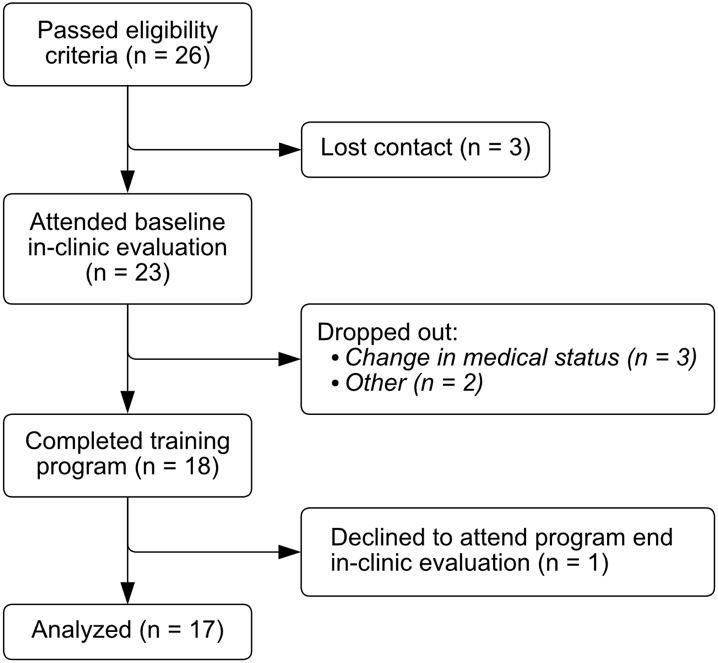

**Figure S1. Patient enrollment.** Flow of enrolled patients through the program.

**Figure S2. Qualitative exit survey results.** Results from the exit survey (n = 19 respondents) about (**A**) satisfaction; (**B**) motivation; (**C**) changes in physical well-being; (**D**) preference of synchronous telerehabilitation vs. in-person sessions; (**E**) participation in additional training; (**F**) ease of fitting the training program into a daily routine; (**G**) support needed during asynchronous training; (**H**) perceived advantage of the home-based training program; and (**I**) patients’  perceived ability to reach their rehabilitation goals by participating in this program.

**Table S1.** **Training movements.** Movements that patients could train with the MindMotion GO and Izar.

| Body region targeted | Movement |
| --- | --- |
| Hand / Wrist | Palmar grasp (grip force) |
|  | Palmar grasping and releasing of the hand (3 activities) |
|  | Finger pinching |
|  | Pincer grasp and release |
|  | Freely squeezing and moving wrist |
|  | Wrist flexion/extension (2 activities) |
|  | Radial/ulnar deviation of wrist |
|  | Forearm pronation/supination (3 activities) |
| Upper Limb | Horizontally reaching with arm |
|  | Vertically reaching with hands |
|  | Reaching with arm and palmar grasp/release |
|  | Unilateral shoulder abduction/adduction |
|  | Bilateral shoulder abduction/adduction |
|  | Horizontal shoulder abduction/adduction |
|  | Pumping movement of arms (bilateral shoulder, elbow, and forearm) |
|  | Shoulder flexion/extension |
|  | Upper limb flexion/extension and lateral flexion of trunk |
| Trunk | Lateral flexion of trunk |
|  | Axial rotation of trunk |
|  | Forward and backwards lateral flexion of trunk |
| Lower Limb | Laterally transferring body weight |
|  | Unilateral stepping |
|  | Bilateral stepping |
|  | Stepping forwards and backwards |
|  | Freely stepping |
|  | Reactive stepping |
|  | Squatting |

**Table S2. Clinical outcomes.** Statistical significance is presented as uncorrected values. Asterisks indicate significance after Bonferroni correction (for 12 comparisons). Abbreviations: FM-UE = Fugl-Meyer Upper Extremities assessment; BBS = Berg Balance Scale; FGA = Functional Gait Assessment; ARAT = Action Research Arm Test; 5xStS = 5 times Stand to Sit; 6minWT = 6-minute Walk Test; TUG = Timed Up and Go Test; HR = Heart Rate; BP = Blood Pressure; NIHSS = National Institutes of Health Stroke Scale; BI = Barthel Index.

|  |  |  | **Potential range** | **Eval. at program start** | | **Eval. at program end** | | **Δ score** | | |
| --- | --- | --- | --- | --- | --- | --- | --- | --- | --- | --- |
| **Assessment** | | ***n*** | **min - max** | **mean (sd)** | **range** | **mean (sd)** | **range** | **mean (sd)** | ***t*** | ***p*** |
| Functional | FM-UE | 17 | 0 - 66 | 31.71 (21.22) | 8 - 65 | 38.12 (20.03) | 14 - 66 | +6.41 (5.09) | 5.20 | 8.8e-5* |
|  | ARAT | 17 | 0 - 57 | 19.00 (21.45) | 0 - 57 | 21.53 (23.29) | 1 - 57 | +2.53 (4.20) | 2.48 | 2.5e-2 |
|  | BBS | 15 | 0 - 56 | 38.67 (11.18) | 14 - 55 | 44.73 (9.15) | 20 - 56 | +6.07 (4.43) | 5.30 | 1.1e-4* |
|  | FGA | 15 | 0 - 30 | 9.87 (6.03) | 1 - 23 | 12.93 (6.20) | 1 - 28 | +3.07 (2.55) | 4.66 | 3.7e-4* |
|  | 5xStS | 15 | 0 - | 22.00 (10.89) sec | 8 - 45 | 17.17 (7.21) sec | 8 - 34 | -4.83 (6.25) sec | -3.00 | 9.6e-3 |
|  | 6minWT | 14 | 0 - | 579.64 (324.83) ft | 120 -1175 | 635.00 (371.46) ft | 130 - 1420 | +55.36 (86.77) ft | 2.39 | 3.3e-2 |
|  | TUG | 15 | 0 - | 31.47 (24.62) sec | 6 - 80 | 27.53 (21.33) sec | 6 - 88 | -3.93 (10.60) sec | -1.44 | 1.7e-1 |
| Physiological | HR | 16 | - | 78.38 (11.22) bpm | 67 - 107 | 72.81 (10.87) bpm | 58 - 96 | -5.56 (10.3) bpm | -2.17 | 4.7e-2 |
|  | BP (systolic) | 16 | - | 114.25 (13.24) mmHg | 88 - 144 | 113.31 (13.88) mmHg | 95 - 148 | -0.94 (13.26) mmHg | -0.28 | 7.8e-1 |
|  | BP (diastolic) | 16 | - | 79.13 (9.70) mmHg | 64 - 92 | 74.44 (7.36) mmHg | 62 - 90 | -4.69 (8.27) mmHg | -2.27 | 3.8e-2 |
| Neuro-logical | NIHSS | 17 | 0 / 42 | 5.41 (3.45) | 1 - 11 | 4.29 (2.91) | 1 - 11 | -1.12 (1.76) | -2.61 | 1.9e-2 |
|  | BI | 17 | 0 / 100 | 85.59 (22.70) | 15 -100 | 87.94 (21.44) | 15 -100 | +2.35 (5.04) | 1.93 | 7.2e-2 |

**Table S3. Patient reported outcomes.** Abbreviations: GAD-7 = General Anxiety Disorder-7; ISI = Insomnia Severity Index; BRS = Brief Resilience Scale; PROMIS = Patient-reported outcomes measurement information system; SIS = Stroke Impact Scale; EQ-5D-5L = European Quality of Life 5 Dimensions 5 Level Version; PHQ-9 = Patient Health Questionnaire.

|  |  | **Potential range** | **Eval. at start** | ***Eval. at end*** | **Δ score** |
| --- | --- | --- | --- | --- | --- |
| **Assessment** | ***n*** | **min - max** | **mean (sd)** | **mean (sd)** | **mean (sd)** |
| GAD-7 | 13 | 0 / 21 | 3.8 (4.2) | 2.0 (2.3) | -1.8* (2.6) |
| ISI | 13 | 0 / 28 | 3.9 (3.5) | 3.3 (3.4) | -0.5 (2.9) |
| BRS | 15 | 1 / 6 | 3.6 (0.9) | 3.8 (0.8) | +0.2 (0.7) |
| PROMIS (social) | 13 | 8 / 40 | 14.4 (6.4) | 14.3 (6.8) | -0.1 (4.1) |
| PROMIS (pain) | 13 | 8 / 40 | 13.3 (5.5) | 13.2 (6.5) | -0.1 (7.6) |
| SIS (strength) | 12 | 0 / 100 | 38.0 (12.1) | 38.0 (17.8) | 0.0 (16.2) |
| SIS (hand) | 12 | 0 / 100 | 17.9 (33.3) | 20.4 (36.3) | 2.5 (10.1) |
| SIS (ADL) | 10 | 0 / 100 | 51.3 (16.4) | 51.8 (19.2) | 0.5 (9.1) |
| SIS (mobility) | 11 | 0 / 100 | 60.4 (28.2) | 67.7 (25.5) | 7.3 (14.0) |
| SIS (communication) | 11 | 0 / 100 | 80.5 (23.1) | 84.1 (20.2) | 3.6 (13.0) |
| SIS (emotion) | 10 | 0 / 100 | 81.9 (17.6) | 83.8 (10.6) | 1.9 (8.9) |
| SIS (memory) | 10 | 0 / 100 | 81.1 (21.5) | 81.8 (20.6) | 0.7 (11.5) |
| SIS (participation) | 9 | 0 / 100 | 42.7 (26.8) | 55.2 (26.5) | 12.5 (18.5) |
| EQ-5D-5L (mobility) | 13 | 1 / 5 | 3.0 (0.9) | 2.8 (0.9) | -0.2 (0.6) |
| EQ-5D-5L (activities) | 13 | 1 / 5 | 3.2 (1.1) | 3.2 (1.2) | -0.1 (0.5) |
| EQ-5D-5L (anxiety) | 13 | 1 / 5 | 1.8 (0.9) | 1.6 (0.7) | -0.2 (1.1) |
| EQ-5D-5L (selfcare) | 13 | 1 / 5 | 2.4 (1.3) | 2.5 (1.2) | 0.1 (0.8) |
| EQ-5D-5L (pain) | 13 | 1 / 5 | 2.1 (0.9) | 1.9 (0.6) | -0.2 (0.7) |
| EQ-5D-5L (vas) | 13 | 1 / 100 | 61.9 (25.8) | 58.8 (20.0) | -3.2 (24.9) |
| SIS (recovery) | 12 | 1 / 100 | 50.8 (20.7) | 49.2 (13.8) | -1.7 (19.5) |
| PHQ-9 | 13 | 0 / 27 | 3.8 (4.2) | 3.9 (3.5) | +0.2 (2.8) |
